# Enhanced Neoplasia Detection in Chronic ulcerative colitis: results of the ENDCaP-C Diagnostic accuracy study

**DOI:** 10.1101/2022.12.15.22283514

**Authors:** The ENDCaP-C Collaborative Group, Ashish Awasthi, Jamie Barbour, Andrew D. Beggs, Pradeep Bhandari, Daniel Blakeway, Matthew Brookes, James Brown, Matthew Brown, Germaine Caldwell, Samuel Clokie, Ben Colleypriest, Abby Conlin, Shanika de Silva, John de Caestecker, Jonathan Deeks, Anjan Dhar, Mark P Dilworth, Edward Fogden, Stephen Foley, Deb Ghosh, Leonie Grellier, Ailsa Hart, Syed Samiul Hoque, Marietta Iacucci, Tariq Iqbal, Jonathan James, Mark Jarvis, Anthoor Jayaprakash, Satish Keshav, Laura Magill, Glenn Matthews, Joel Mawdsley, Simon McLaughlin, Samir Mehta, Kevin Monahan, Dion Morton, Senthil Murugesan, Miles Parkes, Valerie Pestinger, Chris Probert, Arvind Ramadas, Alessandro Rettino, Shaji Sebastian, Naveen Sharma, Michael Griffiths, Joanne Stockton, Venkat Subramanian, Nigel Suggett, Philippe Taniere, Julian Teare, Ajay M Verma, Yvonne Wallis

## Abstract

**Background:** Chronic ulcerative colitis is an inflammatory condition associated with a pro-neoplastic drive, predisposing to colorectal cancer. Repeated colonoscopy is undertaken to detect preneoplastic change, but cancer diagnosis is still frequently missed.

**Aims:** To determine if a predetermined panel of methylation markers could better risk stratify patients, aiding earlier detection of neoplasia.

**Methods:** ENDCaP-C (https://doi.org/10.1186/ISRCTN81826545) was a prospective multicentre test accuracy study of enhanced large bowel neoplasia detection and cancer prevention in patients with chronic ulcerative colitisAll patients underwent baseline colonoscopy and biopsies that had (on central review) shown no dysplasia on histology were put forward for methylation testing. In a prespecified subgroup of 200 patients without initial dysplasia detection, a second colonoscopy was performed, after 12 months.

**Results:** 818 patients underwent a baseline colonoscopy. The methylation assay at baseline (testing <u>non-neoplastic</u> mucosa) was compared with pathology assessment at baseline for neoplasia and showed a diagnostic odds ratio (DOR) of 2.37 (95% CI 1.46, 3.82, P=0.0002). Biopsy analysis was successful in 95% of patients within a multisite routine surveillance programme. The probability of dysplasia increased from 11.1% to 17.7% (13.0%, 23.2%) with a positive methylation result consistent with added value in neoplasia detection.

To determine added value above ‘colonoscopy alone’, a second (reference) colonoscopy was performed in 193 patients without neoplasia. This test also showed an increased number of patients harbouring neoplasia but failed to reach statistical significance (DOR=1.50; 95% CI (0.48, 4.45) P=0.45) The results were also non-significant in the per protocol analysis (DOR=3.93; 95% CI (0.82, 24.75) P=0.09).

Patients with persistent abnormal methylation findings at both colonoscopies were at further enhanced risk of neoplasia, 22% of cases (4/18), or 3x that of patients without methylation changes (7/98).

**Conclusion:** This methylation assay was successfully applied within a routine clinical surveillance programme. Blinded analysis confirmed improved rates of neoplasia detection. Although predetermined levels of statistical significance were not reached, the study has also shown that methylation testing can supplement existing clinical and pathological risk stratification, informing patient consent and anticipated dysplasia detection rates. Although not yet recommended for routine uptake, the finding suggest refined methylation assays could be applied for patient benefit.

## INTRODUCTION

Over half a million patients in the USA are currently affected by ulcerative colitis (UC), a chronic inflammatory condition associated with a pro-neoplastic drive for the development of colorectal bowel cancer(1). The longer the duration of colitis, the more extensive the inflammation, the higher the risk of colorectal cancer (2). The risk of cancer is greatest in those who have been diagnosed young and with extensive colonic inflammation; reaching 18% life time risk after 30 years (3). This results in over 1000 colectomies per year in the UK for colitis associated colorectal cancer or for those in whom (precancerous) dysplastic lesions have been identified (4).

It is likely that neoplastic progression is accelerated by the inflammatory process in UC patients, reducing the ‘window of opportunity’ for early colonoscopic detection of precancerous dysplasia. Despite intensive colonoscopic surveillance with histological assessment of mucosal biopsies, most early tumours are missed. In a large series from the USA, one in six patients had occult invasive cancer in the resected large bowel, highlighting current diagnostic limitations (5). As many as 50% of cases progress to invasive cancer before neoplasia is detected (6, 7) resulting in incurable disease in up to 40% of patients with colitis-associated large bowel cancer (8). Unfortunately, these poor outcomes from surveillance can have an adverse effect upon compliance to repeated colonoscopy (9, 10).

Surveillance programs currently risk stratify patient according to clinical criteria. Accuracy is poor, so international recommendations vary. Current European Society for Gastrointestinal Endoscopy (ESGE) guidelines recommend routine use of pancolonic chromo-endoscopy with targeted biopsies for neoplasia detection in UC (11). Similar guidance has been published by the international SCENIC group (12) who recommend pan-colonic chromo-endoscopy with high-definition endoscopes. A randomized trial (13) has however shown no additional benefit for chromo-endoscopy over HD (high definition) alone, highlighting uncertainty around the best application of these evolving technologies.

To date, colonoscopy-based cancer surveillance programmes have proven inadequate in reliably preventing colorectal cancer in patients with chronic ulcerative colitis. Colonoscopy alone lacks sensitivity in detecting early neoplasia in the presence of inflammatory change in the background large bowel mucosa. Adenomas are a valuable biomarker for colorectal cancer risk in sporadic disease and their removal prevents subsequent cancer development (14), but no comparable benefit has been demonstrated in UCAD. Consequently, when early neoplasia is detected in the presence of UC, although the subsequent cancer risk is low if the lesion can be completely removed endoscopically (15), uncertainty about multifocal disease and compliance to intense follow up prompts clinicians to advise for prophylactic radical resection of the whole colon and rectum; representing overtreatment for many patients and compromising the quality of life for thousands of patients across Europe every year. It remains the case that enhanced neoplasia detection has not translated into reduction in colorectal cancer mortality in patients with longstanding UC nor in the risk of developing interval cancer (16). Better risk stratification through the development of appropriate biomarkers could support endoscopists in providing a more effective and informative surveillance programme, delivering more personalised care with the potential to support the safe introduction of selective organ preserving surgery.

UCAD is associated with a field change predisposition to CRC throughout the damaged mucosa (17). Mucosal molecular biomarkers associated with the earliest stages of neoplastic change can be identified from random (apparently normal) biopsies with the potential to complement colonoscopy by enabling individual patient risk stratification for subsequent cancer development.

DNA methylation promotes tumour suppressor gene silencing or oncogene activation and provides an attractive biomarker for early detection of neoplasia, especially due to the field change phenomenon whereby genetic change can be detected within an organ before histological change. It is known to occur at the early stages of carcinogenesis (18) and can be reliably analysed in DNA extracted from routinely collected formalin fixed paraffin embedded biopsies. We (19) have previously demonstrated tumour associated methylation changes, associated with the earliest (pre-cancer) stages of large bowel tumourigenesis. These epigenetic changes are also seen in pre-cancerous lesions associated with ulcerative colitis (20). Most importantly, these methylation changes have also been detected in the background <u>non-neoplastic</u> mucosa distant from the tumour (21). This field change in the bowel mucosa makes mucosal methylation changes an attractive biomarker for CAD. For the ENDCAP-C trial, five different gene promotor regions were selected from a panel of genes, by analysis of a large multicentre tissue panel (20).

Detecting these methylation changes in the wider field of background mucosa could help to risk-stratify UC patients for neoplastic change, supplementing clinical risk factors and helping prioritise patients for colonoscopic surveillance. Epigenetic silencing of key genes therefore provides both a marker for the development of early (pre cancer) neoplasia and the opportunity to enhance early tumour detection by colonoscopy. Early detection also provides the opportunity to provide local control and avoid radical surgery for a proportion of patients.

We therefore present the results of the ENDCaP-C study is a prospective, multicentre diagnostic accuracy study embedded into a pre-existing surveillance programme for patients undergoing investigation for colitis associated neoplasia. Novel epigenetic markers, derived from DNA extracted from routinely collected bowel mucosal biopsies, were assessed as an adjunctive test to colonoscopy in better risk stratifying patients following examination.

## Methods

### Trial design

The ENDCaP-C trial (https://doi.org/10.1186/ISRCTN81826545) was a prospective multicentre test accuracy study of enhanced large bowel neoplasia detection and cancer prevention in patients with chronic ulcerative colitis. Ethics approval for ENDCaP-C was granted by South East Coast – Surrey Research Ethics Committees (reference 14/LO/1842).

Of 818 recruited patients, 814 underwent baseline colonoscopy, which assessed baseline histology and baseline methylation status. Baseline histology results were made available to patients and clinicians and were used to inform their immediate health care. Methylation test results were not released as these were the index test under evaluation in the study. Those found to be histology negative at baseline were considered to be at risk of having dysplasia missed by colonoscopy and formed the key group of interest in the study to assess whether or not the methylation test can identify cases missed by colonoscopy.

The study sought to recruit 1,000 patients with histologically proven chronic Ulcerative Colitis who were already enrolled on a surveillance programme because of colon cancer risk, and were willing to accept the possibility of an additional colonoscopy between 4 and 12 months after registration. Patients with a history of colorectal cancer were excluded. Eligible participants were identified by review of local IBD databases, in clinic or from endoscopy lists (varied from hospital to hospital). Written informed consent to participate in the study was obtained before registration. All patients underwent baseline colonoscopy by a named colonoscopist as per usual NHS care and routine mucosal biopsy samples were taken.

Endoscopists were permitted to take random biopsies or targeted biopsies, as per their routine practice. The minimum study requirements were two biopsies from the left side of the colon, two from the right side of the colon and one from the rectum. Only biopsies that had shown no dysplasia on histology were put forward for methylation testing.

In the second stage, a reference colonoscopy was undertaken 4–12 months after the baseline colonoscopy to identify dysplasia missed at baseline. Participants were selected for invitation to the second stage if they had no evidence of dysplasia at their index colonoscopy and were undertaken blinded to information from the baseline methylation tests. The histological assessments made from the biopsy samples taken at the second colonoscopy form the reference standard in the study, with which the baseline methylation results were compared. The methylation tests were also repeated in these patients. Biopsy samples were fixed in formalin, embedded in paraffin and processed, and the sections were assessed as per local practice. Analysis of FFPE sections from biopsy material was co-ordinated by a named lead pathologist at each site.

### Methylation index test

The index test was the preselected DNA methylation panel of markers (20). This was tested on the biopsies taken at the baseline colonoscopy. After local histological analysis, samples were screened to ensure that no samples with (histological) neoplasia were sent for DNA analysis. The DNA was extracted and underwent methylation analysis by bisulphite treatment and pyrosequencing(20). To ensure that the reference standard colonoscopy was undertaken blinded to the methylation status, only the study statistician was aware of the positive/negative methylation status.

### Reference Colonoscopy

Patients who were histologically negative at baseline with positive methylation status at the standard surveillance colonoscopy (test positives) were invited to undergo repeat colonoscopy along with selected histologically negative patients who had negative methylation status. Matching by date and site was performed to reduce the chance of bias being introduced.

### Sample size

Computation of the sample size was based on data that dysplasia was detected by histology in 4% of patients from a high-risk cohort, an assumption that a further 4% are missed (assuming a detection rate of 50% for routine colonoscopy) of which 50% will be detected by methylation testing (i.e. sensitivity = 50%) and which will give false-positive results in 5% free of dysplasia (i.e. specificity = 95%).

The study was designed to have adequate power to show that the test was discriminatory (measured by having an odds ratio different from 1) and that the positive predictive value was high enough to be useful for identification of the high-risk patients, being at least 15%.In the cohort of 1000, it was estimated that there would be 80 with underlying dysplasia: 40 of these would be detected by histology from the initial biopsies, and 20 of the remaining 40 would be identified by the methylation test. Following the assumptions about test performance, the methylation test would thus give 46 false-positive results (5% of the 920 without dysplasia) giving an expected positive predictive value of 30% (20 out of 66). With the assumed test performance, a sample size of 66 test positives would have 87% power to show (in a one sample test) a positive predictive value of over 15% with p < 0.05.

Additionally, the status of 132 (double) test negatives were to be verified to obtain estimates of the sensitivity and specificity of the test (computed adjusting for the sampling fraction of test negatives). It was expect that these would include three found positive for dysplasia and 129 without dysplasia, which provides > 90% power to show the diagnostic odds ratio is significantly (p < 0.05) different from one. A specificity of 95% would be estimated with a CI of < 4% points.

### Analysis methods

The analysis for the primary outcome estimates the positive and negative predictive values of methylation as a proportion of those methylation positive at the initial colonoscopy that were detected with colitis-associated neoplasia at the reference colonoscopy, and the proportion of methylation negative that were free of colitis-associated neoplasia at the reference colonoscopy, respectively. The overall discriminatory ability of the methylation test was described using an odds ratio (with 95% CI) with statistical significance assessed by computing Fisher’s exact test comparing follow-up histology rates with baseline methylation status. Values for prevalence (at baseline and reference colonoscopy), sensitivity and specificity were estimated correcting for the sampling proportions of those methylation test positive and methylation test negative using the *svy* complex survey commands in Stata® V15.0 (Stata Corp LP, College Station, TX, USA). Cohen’s kappa was used to assess the agreement between the methylation decision rule at baseline and repeat colonoscopy.

## RESULTS

### Patient Population

Recruitment commenced in November 2014 and ended on 31 March 2017 and follow up completed in March 2018, finishing with 818 patients with chronic ulcerative colitis. In 4 patients no colonoscopy was undertaken, and in 9 cases no mucosal biopsies were sent for histology. The remaining 805 patients, selected from 4,800 patients across 31 hospitals underwent an index colonoscopy with multiple mucosal biopsies taken for histology and methylation testing (Figure 1).

**Figure 1:**
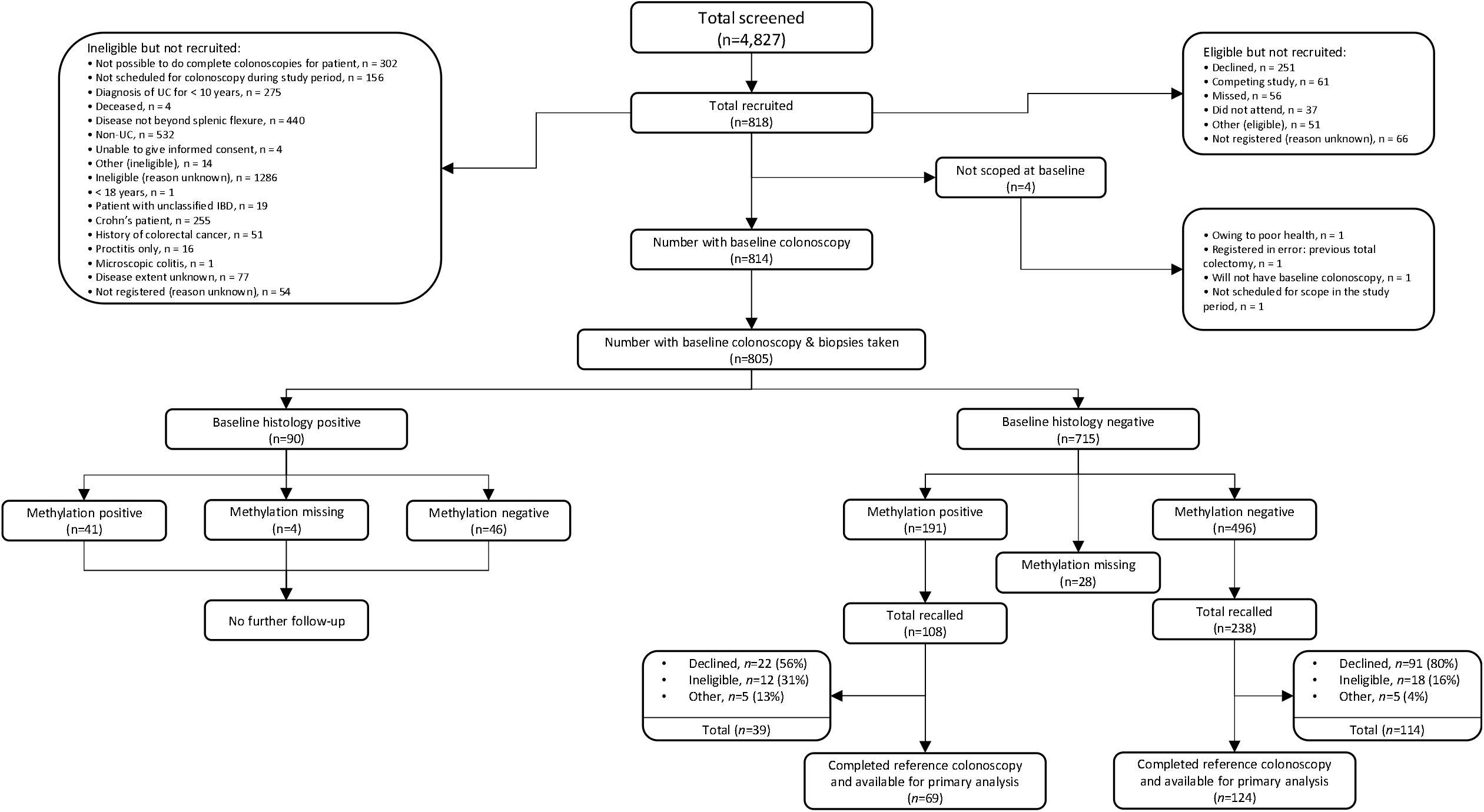
The Standards for Reporting of Diagnostic Accuracy (STARD) flow diagram.

The baseline data for participants are shown in Table 1 and represent a cohort at high risk of colitis associated neoplasia. Additional risk factors included 24.5% of the population suffering from PSC, 72% with disease to the splenic flexure or beyond and a mean age of 53yrs with standard deviation [14yrs].

**Table 1:**
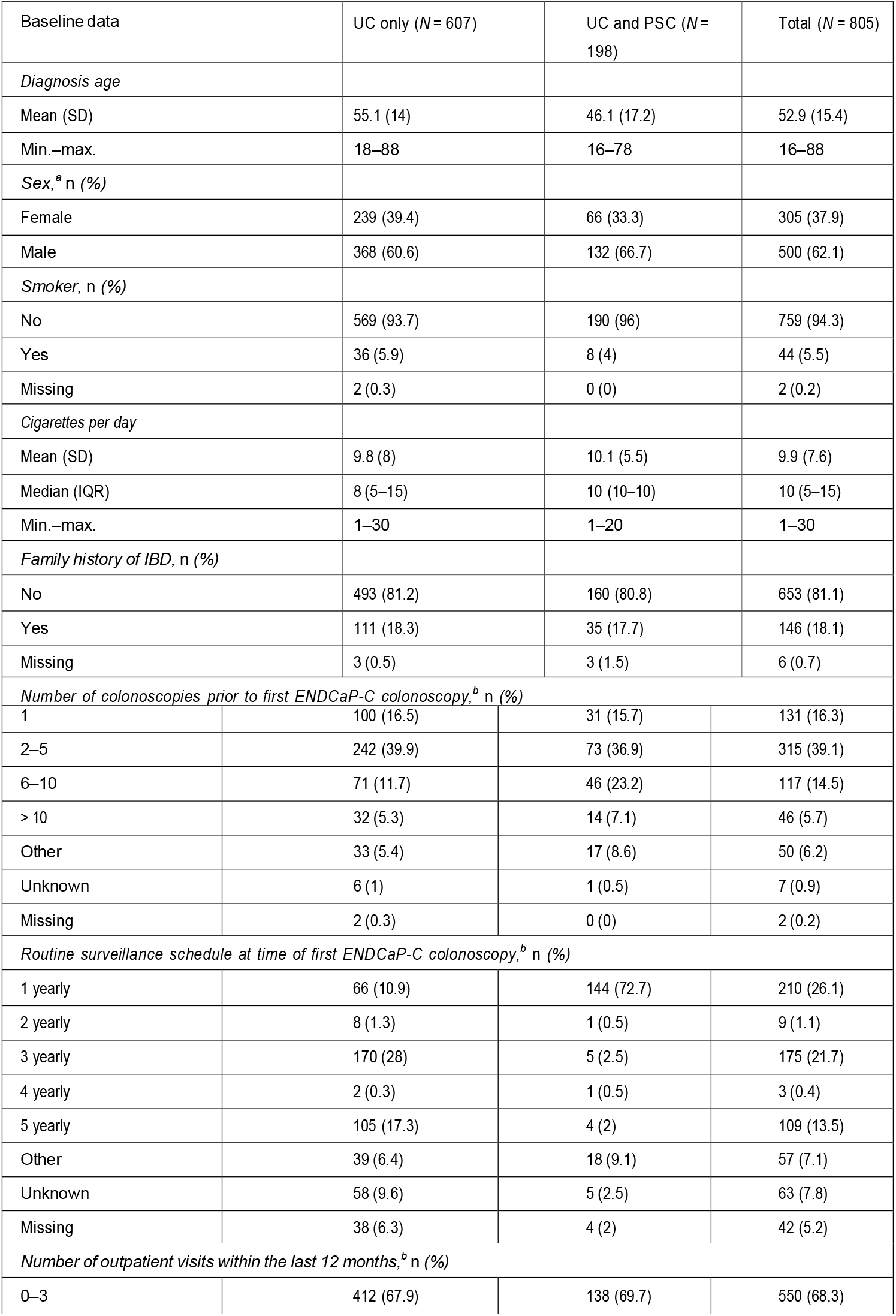
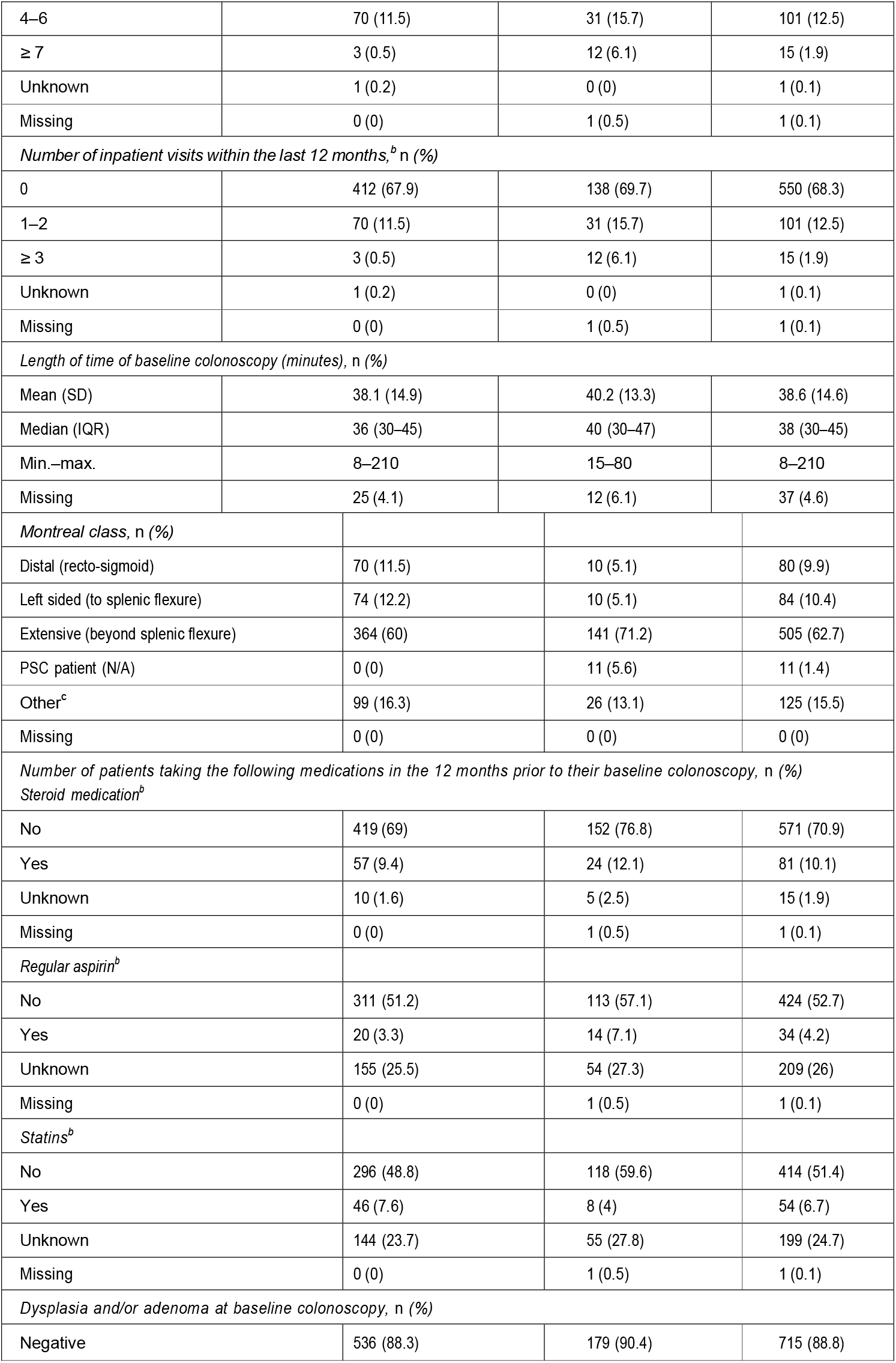
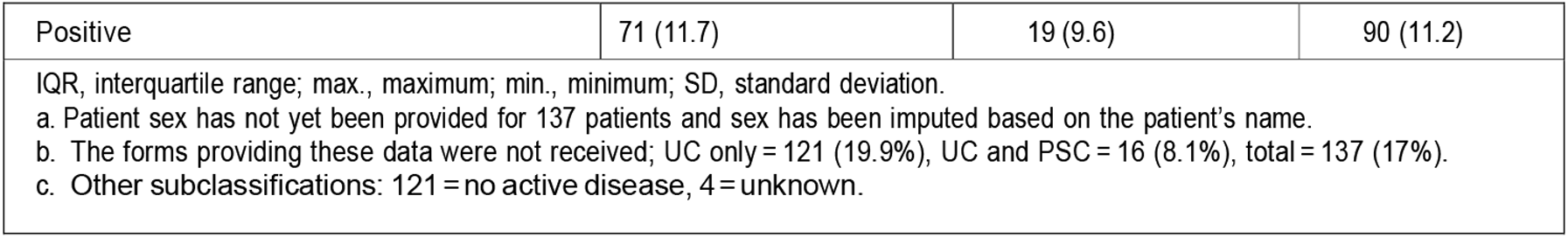
Baseline data of ENDCAP-C trial participants:

### Association of aberrant methylation in non-neoplastic large bowel mucosa with synchronous large bowel neoplasia

Firstly, we wished to understand whether measurement of our pre-specified methylation markers in histologically normal mucosa could be used to predict the concurrent presence of dysplasia or neoplasia elsewhere in the colon.

We found a statistically significant association between methylation test results in histologically normal mucosa and the presence of dysplasia or neoplasia elsewhere in the colon, in the biopsies taken at the reference examination, with a diagnostic odds ratio (DOR) of 2.37 (95% CI 1.46 to 3.82; p < 0.001). Crucially, this effect was sustained where only results from patients with dysplasia were included with a DOR of 2.06 (95% CI 1.28, 3.33; p<0.001). This demonstrates that the background methylation signature is associated with observed concurrent precancerous neoplasia and therefore could help stratify patients as to the risk of underlying neoplasia. This association was independent of the diagnosis of PSC in whom the incidence of neoplasia was 9.6%. The baseline results showed a higher than anticipated baseline prevalence of dysplasia: 11.2% (90/805), consistent with a specially selected high risk population and also indicative of a high standard of examination.

### The Reference Standard Examination (second colonoscopy)

Figure 1 shows the selection of the patients for the reference colonoscopy, which was undertaken in 193 patients (between 4 months and 2 years after the index procedure), all of whom had no evidence of neoplasia at the index colonoscopy procedure. Patients were selected in proportions 2:1 (for the absence of methylation changes: positive methylation testing); excluding those patients with histologically confirmed dysplasia at the index colonoscopy (n=90).

#### Primary analyses

This compared the baseline methylation results with the histology analysis of biopsy samples from the reference colonoscopy.

A total of 193 participants had data available for these analyses, with 172 eligible for the sensitivity analysis, 104 for the per-protocol analysis and all 193 for the intention-to-treat analysis. Table 3 reports the performance of the methylation test for these three pre-planned analyses.

#### Sensitivity analysis

excluding absence of dye spray or non-standard endoscopic technique, 172 of the 193 patients fulfilled these criteria. This analysis standardised the colonoscopy procedure across the participating centres. This was an attempt to quality assure the reference colonoscopy. In this analysis, the association between methylation status and histology findings was in the direction of positive methylation, indicating an increased risk of neoplasia; however, this did not reach statistical significance DOR of 2.01 (95% CI 0.60, 6.84; p = 0.27). Fifteen of the 172 patients in this analysis were histology positive on the reference colonoscopy (8.3%). The methylation test identified eight of these patients (sensitivity of 44%; 95% CI 20% to 72%).

A positive index methylation test increased the probability of identifying dysplasia at the reference examination to 12% (8/65); Of clinical relevance, for those patients with a +ve methylation test, a ‘normal’ index colonoscopy did not reduce the risk of neoplasia at the (planned 1 year) follow up period. By contrast, a methylation negative result reduced by 40%, the risk of neoplasia during the follow up period (6.5%; 7/107 reference examinations).

#### Per-protocol analysis

104 patients were eligible for this analysis. It excluded 68 patients who did not receive their reference colonoscopy within 12 months of the baseline colonoscopy. This reduces the potential for de novo dysplasia being identified at the reference examination (stringency test). Diagnostic odds ratios was increased to around 4 but still did not reach levels of conventional statistical significance DOR of 3.93 (95% CI 0.82, 24.8; p = 0.09). A positive baseline methylation test result (despite no neoplastic biopsies), increased the probability of identifiable colonic neoplasia within 1 year to 17%. Being methylation negative reduced the prevalence of neoplasia within the follow up period to under 5% (3/62 examinations).

#### The intention-to-treat analysis

including all patients, found weaker discrimination with diagnostic odds ratios of 1.50 (95% CI 0.48, 4.45; p = 0.45). A positive methylation test result still increased the probability of being histology positive within the follow up period to 12%. Whilst being methylation negative reduced the probability of being histology negative to 8%.

## CONCLUSIONS

This is the first prospective multicentre trial to report on mucosal methylation as a biomarker of neoplastic change in the colon. The study has evaluated a pre-selected methylation signature in a ‘real world’ surveillance setting across 31 hospitals in the UK. The study has shown a clear and statistically significant association between defined promoter methylation changes in background colonic mucosa and histological evidence of distant neoplastic change in the large bowel, strongly supporting the underlying hypothesis. The second (reference) colonoscopy, looking for ‘missed neoplasia’, demonstrated the same association between the initial methylation findings and neoplasia detection, but the association was weaker and statistically non-significant. The potential clinical value of this approach was demonstrated, with improved stratification of patients for both high and for low risk of concurrent neoplasia.

ENDCAP was embedded within the existing surveillance programme for UC across 30 different NHS hospitals. Histological assessment and methylation testing was performed within routine NHS laboratory facilities. We demonstrated that the additional test was acceptable to patients and clinicians and could be incorporated within existing care pathways at 10 different hospitals.

Quality assurance is challenging within any pragmatic multicentre study. The high incidence of neoplasia (11%) at the index (routine) colonoscopy was reassuring. The compliance to dye spray (90%) examination at the reference test was also higher than we anticipated. Ultimately we prioritised real world testing, to evaluate the practicalities of delivering such a test. The one significant protocol violation was the delay beyond 12 months for the reference examination, often reflecting the capacity pressures within the NHS endoscopy service. We were however able to undertake a per-protocol analysis to account for the impact of de-novo neoplasia and other confounding variables that may have been influenced by the delay.

There was a substantial rising incidence of neoplasia beyond the 12-month window particularly seen in the control arm of the reference examination, perhaps related to selection bias in recalling the control group. It was seen that the risk of patients declining repeated examination was almost 2x higher in the control arm as compared to the methylation positive group (20% vs 38%), raising the possibility that a higher risk population was being (subconsciously) selected. The methylation results were blinded to all clinical trials staff, clinicians, and patients, but the patient and physician could not be blinded as to the macroscopic colonoscopy findings; this information may have influenced decision making.

The study population was selected to be at high risk of neoplasia as defined by the associated risk factors seen in the study population. The proportion of patients with identified dysplasia (11.2%) at the baseline colonoscopy was considerably higher than predicted and that has been seen in other studies (22). The high prevalence was not explained by inclusion of PSC patients as their observed dysplasia rate was lower, presumably due to being a younger cohort.

Methylation changes were seen in only 50% of patients harbouring neoplastic changes at index colonoscopy. We selected the methylation targets from a large multicentre retrospective cohort (Beggs). It may be that testing a wider range of methylation sites and repeated testing would increase the sensitivity of the test.

At reference colonoscopy, we were able to assess the impact of repeat methylation testing. The numbers were necessarily small, but did show evidence of additional risk stratification; where repeated positive methylation tests were associated with a rising risk of neoplastic change. As methylation biomarkers are further investigated, these results will be clarified.

It is plausible that a subset of CUC patients will harbour dysplasia without manifesting a methylation field change and our results would support this. Almost half the patients with dysplasia had no methylation changes at baseline or at follow up examination. It is rational to propose such patients represent a lower risk population, perhaps even sporadic disease (rather than colitis driven dysplasia). If this is the case, these patients would be suitable for local treatment of dysplasia and so avoid radical surgery. This is a current and pressing clinical issue that methylation testing of the background mucosa could help address. Longitudinal follow-up of the ENDCaP-C patient population will assess residual neoplasia risk and further inform on the predictive value of background mucosa methylation changes.

For patients who have positive methylation changes in their background mucosa, but no dysplasia-ENDCAP indicates their risk of developing dysplasia in the following 12 months (per protocol analysis) is higher than before their ‘*normal*’ index colonoscopy (17% in the next 12 months), despite the initial ‘reassuring’ colonoscopy findings. On the other hand, for a similar patient without evidence of mucosal methylation change, the chance of dysplastic change being identified at colonoscopy in the next 12 months is reduced to 3/62 (∼5%).

Importantly, the proportion of patients identified at higher risk is less than 1 in 3 (191/715). The majority, 73% have more than halved their risk of manifesting dysplasia in the following 12 months. For patients with repeated positive methylation signatures, their risk of neoplasia being identified within 12 months is over 20%.

It would not be appropriate, and we would not recommend using methylation testing in isolation, to guide clinical decision making. However in combination with high quality endoscopic examination, better risk stratification could be achievable. These two high risk subgroups could be prioritised for surveillance and may benefit from more frequent colonoscopies. Informed decision making is helpful to both clinicians and patients and may benefit adherence to surveillance colonoscopy.

Clinical stratification is currently rather crude. It does not use ‘normal’ endoscopic findings to modify an individual’s risk. Molecular stratification can complement/inform the surveillance programme to increase frequency of examination for high risk patients and help reassure the patient and endoscopist of low risk status.

Stratification might also differentiate between multifocal and localised disease. 40% of dysplasia showed no background changes. These may define a group more suitable for local excision.

Methylation testing can improved risk stratification. This might help compliance which is currently low in this patient population by reassuring endoscopists and encouraging patients to undergo repeated examinations.

## Supporting information

STARD Checklist

## Data Availability

Data not available as confidential health data

## APPENDIX

ENDCAP-C Collaborative:

The principal investigator at each centre is indicated with an asterisk:

**Bart’s Health NHS Trust**

Konstantinos Giaslakiotis, Ceri Guarnieri, Syed Samiul Hoque,* Bianca Petri and Attia Rehman.

**Basildon and Thurrock University Hospitals NHS Foundation Trust**

Nazar Alsanjari, Wilson Alvares, Georgina Butt, Natasha Christmas, Mark Jarvis,* Denis Lindo, Zia Mazhar, Pushpakaran Munuswamy, Javaid Subhani and Gavin Wright.

**Blackpool Teaching Hospitals NHS Foundation Trust**

Senthil Murugesan,* Greta Van Duyvenvoorde, Suboda Weerasinghe and Rachael Wheeldon.

**Cambridge University Hospitals NHS Foundation Trust**

James Yiu Hon Chan, Miles Parkes,* Konstantina Strongili and Merry Jay Jimenez-Smith.

**County Durham and Darlington NHS Foundation Trust**

Sarah Clark, Anjan Dhar,* Gillian Horner and Sree Mussunoor.

**Gateshead Health NHS Foundation Trust**

Jamie Barbour,* Sophie Gelder, James A Henry, Paul O’Loughlin, Jitendra Singh and Ann Wilson.

**Hampshire Hospitals NHS Foundation Trust**

Matthew Brown,* Susan Farmer, Asmat Mustajab, Caroline Palmer and John Ramage.

**Heart of England NHS Foundation Trust**

Safia Begum, Gerald Langman and Naveen Sharma.*

**Hull and East Yorkshire Hospitals NHS Trust**

Sarah Ford, Laszlo Karsai, Sally Myers and Shaji Sebastian.*

**Imperial College Healthcare NHS Trust**

Robert Goldin, Bridget Hradsky, Christwishes Makahamadze, Mike Osborn, Simon Peake and Julian Teare.*

**Isle of Wight NHS Trust**

Leonie Grellier,* Kamarul Jamil, Andres Kulla, Norman Mounter, Chris Sheen and Joy Wilkins.

**Kettering General Hospital NHS Foundation Trust**

Laura Aitken and Ajay Verma.*

**Leeds Teaching Hospitals NHS Trust**

Suzie Colquhoun, Ruth Fazakerley, Olorunda Rotimi, Venkat Subramanian* and Charlotte Wilson.

**London North West Healthcare NHS Trust**

Ibrahim Al-Bakir, Naila Arebi, Pooja Datt, Ailsa Hart,* Dennis Lim, Ravi Misra, Morgan Moorghen and Lawrence Penez.

**Northumbria Healthcare NHS Foundation Trust**

Shoba Abraham, Helen Bailey, Sonya Beaty, Jane Dickson, Anthoor Jayaprakash* and Linda Patterson.

**Oxford University Hospitals NHS Trust**

Nathan Atkinson, Amirah Bouraba, James East, Satish Keshav,* Conor Lahiff, Mary Lucas, Simon Travis, Lai Mun Wang, Kate Williamson and Jean Wilson.

**Portsmouth Hospitals NHS Trust**

Michelle Baker-Moffatt, Pradeep Bhandari,* Jennifer Hale, Maria Hayes, Kesavan Kandiah, Lisa Murray, David Poller, Sharmila Subramaniam, Sreedhari Thayalasekaran and Prokopios Vogiatzis.

**Royal Liverpool and Broadgreen University Hospitals NHS Trust**

Tim Andrews, Fiona Campbell, Martyn Dibb, Tracey Forshaw, Alvyda Gureviciute, Hayley Hamlett, Chris Probert,* Sreedhar Subramanian and Clare Shaw.

**Royal United Hospital Bath NHS Foundation Trust**

Leigh Biddlestone, Dawne Chandler, Ben Colleypriest* and Lucy Howie, Joyce Katebe.

**Salford Royal NHS Foundation Trust**

Gordon Armstrong, Abby Conlin* and Melanie Taylor.

**Sandwell and West Birmingham Hospitals NHS Trust**

Ala Ali, Julie Colley, Edward Fogden,* Ann Francis, Anne Hayes, Samia Hussain, Imtiyaz Mohammed, Suhail Muzaffar, Nithin Nair, Mirza Sharjil Baig and Nigel Trudgill.

**Sherwood Forest Hospitals NHS Foundation Trust**

Stephen Foley,* Cheryl Heeley, Samiya Ibrahim and Wayne Lovegrove.

**South Tees Hospitals NHS Foundation Trust**

Andrea Boyce, Wendy Jackson, David Oliver, Kolanu Prasad, Arvind Ramadas* and Julie Tregonning.

**The Dudley Group NHS Foundation Trust**

Clare Allcock, Shanika de Silva,* Sarah Fullwood, Shahkee Garai, Sauid Ishaq, Rizwan Mahmood, Ali Sherif, Veena Shinde and Claire Whitcombe.

**The Princess Alexandra Hospital NHS Trust**

Deb Ghosh,* Hazel Guth, Nikki Staines and Vasi Sundaresan.

**The Royal Bournemouth and Christchurch Hospitals NHS Foundation Trust**

Nina Barratt, Emma Gunter, Joan McCutcheon and Simon McLaughlin.*

**The Royal Wolverhampton NHS Trust**

Matthew Brookes,* Jill Brown, Dragana Cvijan, Marie Green, Shingankutti A M Mangalika, Jayne Rankin, Karen Sedgewick, Asit Shah and Monika Widlak.

**University Hospitals Birmingham NHS Foundation Trust**

Olufunso Adedeji, Claire Arthur, Lillie Bennett, Neeraj Bhala, Louise Bowlas, Ralph Boulton, Ruth Buckingham, Tamar Cameron, Rachel Cooney, Sheldon Cooper, Christopher Dobson, Jason Goh, Tariq Iqbal,* Kate Kane, Shrikanth Pathmakanthan, Zarqa Shoaib, Jessica Simmons, Nigel Suggett, Philippe Taniere, Tanya Van der Westhuizen and Robert Walt.

**University Hospitals of Leicester NHS Trust**

John de Caestecker,* Angus McGregor, Alison Moore and Sarah Nicholson.

**Walsall Healthcare NHS Trust**

Katherine Johanna Arndtz, Ashish Awasthi,* Kiran Desai, Victoria Foster, Rachael Howe, Rangarajan Kasturi and Jasbir Nahal.

**West Middlesex University Hospital NHS Trust**

Shaila Desai, Joel Mawdsley,* Kevin Monahan,* Metod Oblak, Krishna Sundaram and Anne Thorpe.

